# Analysis of C-reactive protein omics-measures associates methylation risk score with sleep health and related health outcomes

**DOI:** 10.1101/2024.09.04.24313008

**Authors:** Ziqing Wang, Danielle A Wallace, Brian W Spitzer, Tianyi Huang, Kent Taylor, Jerome I Rotter, Stephen S Rich, Peter Y Liu, Martha L. Daviglus, Lifang Hou, Alberto R Ramos, Sonya Kaur, J Peter Durda, Hector M González, Myriam Fornage, Susan Redline, Carmen R Isasi, Tamar Sofer

## Abstract

**Introduction:** DNA methylation (DNAm) predictors of high sensitivity C-reactive protein (CRP) offer a stable and accurate means of assessing chronic inflammation, bypassing the CRP protein fluctuations secondary to acute illness. Poor sleep health is associated with elevated inflammation (including elevated blood CRP levels) which may explain associations of sleep insufficiency with metabolic, cardiovascular and neurological diseases. Our study aims to characterize the relationships among sleep health phenotypes and CRP markers —blood, genetic, and epigenetic indicators— within the Hispanic Community Health Study/Study of Latinos (HCHS/SOL).

**Methods:** In HCHS/SOL, methylation risk scores (MRS)-CRP and polygenetic risk score (PRS)-CRP were constructed separately as weighted sums of methylation beta values or allele counts, respectively, for each individual. Sleep health phenotypes were measured using self-reported questionnaires and objective measurements. Survey-weighted linear regression established the association between the multiple sleep phenotypes (obstructive sleep apnea (OSA), sleep duration, insomnia and excessive sleepiness symptom), cognitive assessments, diabetes and hypertension with CRP markers while adjusting for age, sex, BMI, study center, and the first five principal components of genetic ancestry in HCHS/SOL.

**Results:** We included 2221 HCHS/SOL participants (age range 37-76 yrs, 65.7% female) in the analysis. Both the MRS-CRP (95% confidence interval (CI): 0.32-0.42, p = 3.3 x 10^-38^) and the PRS-CRP (95% CI: 0.15-0.25, p = 1 x 10^-14^) were associated with blood CRP level. Moreover, MRS-CRP was associated with sleep health phenotypes (OSA, long sleep duration) and related conditions (diabetes and hypertension), while PRS-CRP markers were not associated with these traits. Circulating CRP level was associated with sleep duration and diabetes. Associations between OSA traits and metabolic comorbidities weakened after adjusting for MRS-CRP, most strongly for diabetes, and least for hypertension.

**Conclusions:** MRS-CRP is a promising estimate for systemic and chronic inflammation as reflected by circulating CRP levels, which either mediates or serves as a common cause of the association between sleep phenotypes and related comorbidities, especially in the presence of diabetes.

## Introduction

Sleep disorders are risk factors for impaired neurological function, metabolic and cardiovascular diseases (CVD)^1^. For instance, obstructive sleep apnea (OSA), characterized by episodic upper airflow narrowing or collapse^2^, causes intermittent hypoxia, systemic inflammation and endothelial dysfunction. These physiological disturbances are considered key pathways linking OSA to increased risk for hypertension, cardiovascular and cerebrovascular diseases^3,4^. Sleep fragmentation and restriction-characteristics of many sleep disorders, including insomnia, have also been shown to elevate inflammation level and cardiovascular morbidity^5–7^. Furthermore, excessive daytime sleepiness (EDS) is a symptom of insufficient sleep or heightened sleep propensity and is also found associated with inflammation^8^.

C-reactive protein (CRP), as a blood marker of systemic inflammation, is commonly used as a risk indicator for cardiometabolic diseases, including coronary heart disease, diabetes, and hypertension^9–11^. However, circulating CRP levels fluctuate following acute diseases, diminishing its stability and accuracy in estimating chronic inflammation. While sleep disorders, including OSA, have been associated with systemic inflammation^12^, early studies reported conflicting results on associations of OSA with blood CRP levels, likely due to limitations such as small sample size, study designs overlooking confounding factors including body mass index (BMI), detection threshold of CRP and so on^13,14^.

DNA methylation (DNAm) is an epigenetic modification essential for transcriptional regulation and various fundamental biological processes^15^, impacted by both genetics and environment^16^. While DNAm has site-specific effects, overall DNAm variability over time in adults is relatively stable^17^. Methylation risk scores (MRS) have been developed as weighted sums of DNAm at CpG sites and tested for their associations with health outcomes or biomarkers (such as aging or CRP) to infer exposures and disease states, similar to polygenic risk scores (PRS). Recently, an MRS for CRP (MRS-CRP) has emerged as a more accurate marker for chronic inflammation than circulating CRP, exhibiting enhanced test-retest reliability and stronger associations with long term health outcomes ^18–21^. CRP levels are also partly determined by genetics, with genetic loci associated with CRP explain approximately 10% of variation in blood CRP level^21,22^.

The current study aims to investigate the relationships between circulating, epigenetic and genetic CRP patterns and common sleep problems (OSA; insomnia and EDS; sleep duration); as well as sleep-related comorbidities (hypertension, diabetes and cognitive decline); in the Hispanic Community Health Study/Study of Latinos (HCHS/SOL) cohort. For secondary analysis, we estimated the associations of sleep phenotypes with sleep-associated comorbidities, with and without adjusting for MRS-CRP to assess the extent to which MRS-CRP potentially explains the associations. Additionally, lasso penalized regression was performed to select key CRP-associated CpG sites associated with sleep traits and their comorbidities. The contribution of this paper is two-fold: 1) addressing the knowledge gap regarding the associations between MRS-CRP and sleep traits; 2) exploring associations between CRP omics measures with sleep, metabolic syndrome and cognitive function in a Hispanic population.

## Methods

Figure 1 illustrates the overview of the study design. Since our primary interest is modeling associations in the HCHS/SOL cohort, we first constructed various PRS-CRPs in an independent cohort, the Multi-Ethnic Study of Atherosclerosis (MESA) using multiple methods and summary statistics. The best-performing PRS-CRP, selected based on its association with circulating CRP in MESA, and MRS-CRP were then computed using data from the Hispanic Community Health Study/Study of Latinos (HCHS/SOL) cohort, followed by association tests with blood CRP levels. We evaluated associations of these CRP omics-measures with sleep, cognitive and metaboli comorbidities via survey linear regression, and conducted secondary analyses to further explore the relationships between systemic inflammation and these health conditions from epigenetic and genetic perspectives.

**Figure 1.**
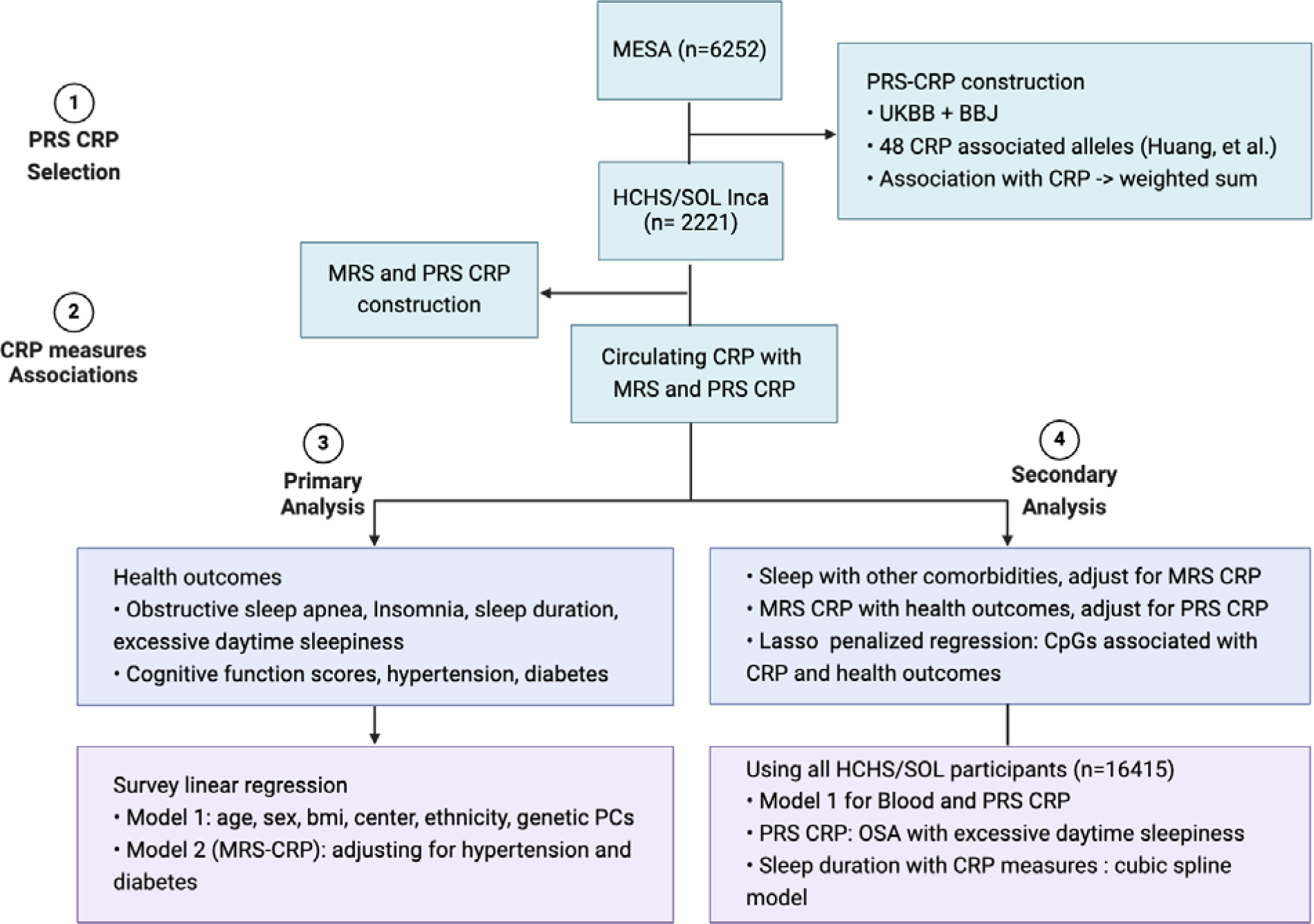
Study design. Best-performing polygenic risk score (PRS) of C-reactive protein (CRP) was first selected in MESA as the one explaining the highest variance in circulating CRP. Then association of circulating CRP with Methylation risk score (MRS) and PRS constructed in SOL-INCA was confirmed before evaluating the association of these CRP measures with sleep and other health outcomes.

### The MESA Dataset

The MESA is a multi-center longitudinal cohort study comprising 6,814 men and women aged 45–84 years, self-identified from four race/ethnic groups without overt clinical CVD at the time of enrollment (Exam 1; 2000-2002)^23^. The current study included all participants with CRP measured at Exam 1 (N=6,415) with appropriate consent and excluded N=163 individuals without genetic data in the current dataset.

#### Assessment of CRP and Demographic Characteristics

High sensitivity CRP was measured during MESA Exam 1 using a BNII nephelometer with particle-enhanced immunonephelometric assays (Dade Behring Inc, Deerfield, IL). Sociodemographic characteristics, including age, sex and medical history, were collected using standard self-reported questionnaires. To report demographic characteristics, participants were stratified by clinical CRP risk groups for CVD (< 1mg/L [low risk], 1–<3 mg/L [Borderline], and ≥ 3mg/L [elevated risk])^24^.

#### Whole-genome sequencing (WGS) and PRS construction

MESA WGS data was obtained from the Trans-Omics in Precision Medicine (TOPMed) program Freeze 10 release, with reads aligned to the human-genome build GRCh38 by a common pipeline across all sequencing centers^25^. Variant and genotype calling was performed jointly on all samples in a given TOPMed freeze^25^. Minor allele frequency (MAF) threshold for variant inclusion was set to MAF > 0.01 (computed over MESA participants) and genetic relatedness was computed centrally by TOPMed using the PC-Relate and algorithm^26^.

Multiple methods were employed to construct genome-wide association analysis (GWAS)-derived PRS-CRP using summary statistics from UK Biobank (UKBB)^27^ and Biobank Japan (BBJ)^28^ for MESA and HCHS/SOL participants. UKBB data were divided into sets representing different genetic backgrounds: AFR (African, n = 6203), AMR (Amerindian, n = 937), EAS (East Asian, n = 2564), EUR (European, n = 400,094), and SAS (South Asian, n = 8397)^27^.

First, UKBB-EAS and BBJ data were meta-analyzed using GWAMA (Genome-Wide Association Meta-Analysis) software^29^ with default parameters allowing indel alleles. The GWAMA output, together with other UKBB ancestry-specific GWAS summary statistics, was then submitted to PRS-CSx^30^ for development of variant weights. In PRS-CSx, variants present in at least one of the UKBB linkage disequilibrium reference panels were used to develop posterior effect estimates for each of the five genetic backgrounds^30^, based on which PRSs were constructed using PRSice2^31^ without clumping or thresholding. We also developed PRSs using PRSice2, based on UKBB+BBJ meta-analyzed GWAS summary statistics, with clumping and thresholding set to R^2^ = 0.1, distance = 1000 Kb, and p-value = 5 x 10^-8^, 1 x 10^-7^, 1 x 10^-6^, 1 x 10^-5^, 1 x 10^-4^, 1 x 10^-3^, 1 x 10^-2^, 0.1.

A simple PRS summing the effect estimates for 48 genome-wide significant alleles associated with CRP^8^ (referred to as Huang et al. PRS) was also constructed using PLINK^32^. For comparison purposes, each calculated PRS was standardized by subtracting its mean and divided by its standard deviation.

#### PRS association analysis with CRP levels in MESA

Association between each PRS-CRP and blood CRP level was assessed using simple linear regression in MESA, while adjusting for age, sex, body mass index (BMI), race, study site and first 5 genetic PCs. The best-performing PRS-CRP was determined as the one explaining highest variance in blood CRP levels and used in subsequent analysis in HCHS/SOL. We also carried forward a PRS developed as a weighted summation of PRS-CSx PRSs with weights determined by the regression coefficients from CRP regressed over all ancestry-specific PRS-CSx PRSs in MESA, where we further used either all MESA individuals, or only MESA Hispanic individuals. We do not report the results from association analyses of this PRS in MESA due to overfitting (as weights are developed based on analysis in MESA), but it was used in HCHS/SOL based on past work that demonstrated that such a weighted sum tends to outperform other PRSs^33^.

#### The HCHS/SOL dataset

The HCHS/SOL is a population-based cohort study with participants of participants who were recruited from four metropolitan areas across the United States (Bronx NY, Chicago IL, Miami FL, and San Diego CA)^34^. By design, participants represented diverse Hispanic/Latino origins (and genetic admixture) and had a wide age range (18-74 years old at baseline). Among the total 16,415 participants, 2,221 from the Study of Latinos-Investigation of Neurocognitive Aging (SOL-INCA) ancillary study were considered for inclusion due to available genetic data and data on methylation, sleep phenotypes and circulating CRP. Sociodemographic and lifestyle variables were self-reported, including age, sex, household income level, smoking status, cigarette and alcohol consumption, as well as self-identified Hispanic background. Obesity parameters BMI and waist-hip ratio were measured following standard procedure at baseline. BMI was calculated as body weight (kg) divided by height squared (m²), and waist-hip ratio as waist circumference divided by hip circumference.

#### Sleep assessment

Multiple sleep phenotypes were assessed using self-reported questionnaires from the baseline visit^35^. Sleep duration was derived as weighted average of weekday and weekend sleep. Long and short sleep variables were derived from sleep duration as binary outcomes, with thresholds set at more than 9 hours and less than 6 hours. Scores indicative of insomnia and excessive daytime sleepiness were also measured using the 5-item Women’s Health Initiative Insomnia Rating Scale (WHIIRS)^36^ and the 8-item Epworth Sleepiness Scale (ESS)^37^, respectively. OSA was assessed using a self-applied sleep apnea monitor (ARES Unicorder 5.2; B-Alert, Carlsbad, CA) that measured nasal airflow, position, snoring, actigraphy, and hemoglobin oxygen saturation (SpO2)^38^ during overnight sleep. Obstructive respiratory events were defined as a 50% or greater reduction in airflow with associated desaturations of 3% or larger, lasting for at least 10 seconds. Apnea–hypopnea index (AHI) was subsequently computed as the sum of such events per estimated sleep hour^35^. Given the importance of measures of hypoxia in characterizing OSA severity^39^, we also studied several measures of overnight desaturation (minimum SpO2, average SpO2 and % Time SpO2<90 - the percentage of cumulative time with oxygen saturation below 90% in total sleep time).

#### Assessment of C-reactive Protein and Relevant Comorbidities

In HCHS/SOL, high sensitivity CRP was assayed with a Roche Modular P Chemistry Analyzer (Roche Diagnostics Corporation, IN) using the immunoturbidimetric method (Roche Diagnostics Corporation, IN) during the baseline visit^40^ (Visit 1). Demographic characteristics of the HCHS/SOL target population were summarized by clinical CRP risk groups for CVD (< 1mg/L [low risk], 1–<3 mg/L [Borderline], and ≥ 3mg/L [elevated risk])^24^.

Diabetes status was identified (Visit 1) based on the American Diabetes Association definition, taking into account fasting serum glucose level (≥126 mg/dL), glucose tolerance test (≥200 mg/dL), medication use and hemoglobin A1c levels (HbA1c, ≥ 6.5%), whichever was available^41^. Hypertension was identified as the presence of either systolic (≥140 mmHg) or diastolic blood pressure (≥90 mmHg) exceeding the threshold, or by the use of antihypertensive medications^41^. Cognitive function score at baseline (Visit 1) was computed as the mean of the standardized scores of three cognitive tests^42^: 1) Brief-Spanish English Verbal Learning Test (B-SEVLT; verbal episodic learning and memory); 2) Word Fluency test (WF; verbal fluency); 3) Digit Symbol Substitution test (DSS; processing speed and executive function)^42^. SOL-INCA ancillary study, conducted concurrently or within a year of Visit 2 (7 years after Visit 1, on average), repeated the same neuropsychological tests with additional two Trail Making Tests (TMT-A and B; executive function), which were also included in the calculation of the cognitive function score^42^. Cognitive change was defined as the difference between the cognitive function score at SOL-INCA and Visit 1.

#### Genotype imputation and PRS c onstruction

HCHS/SOL participants were genotyped on a custom Illumina HumanOmni2.5-8v1-1array with 150,000 added custom variants, including ancestry informative single-nucleotide polymorphism (SNPs)^43^. Over 12,000 samples were imputed using the TOPMed imputation server^43^. As in MESA, MAF threshold was applied for variant exclusion in the SOL-INCA dataset, requiring MAF≥0.01, where MAF was computed in HCHS/SOL. PRS-CRP for the HCHS/SOL dataset used in the study was computed using the same method as the best performing PRS-CRP selected in MESA, and PRS summation of PRS-CSx was computed with weights obtained from regression of CRP on ancestry-specific PRS-CSx PRSs using all MESA individuals and only MESA Hispanic individuals, as described above.

#### Epigenome-wide methylation quantification and MRS construction

Methylation profiling of whole blood samples from SOL-INCA participants across the genome was performed using the Infinium MethylationEPIC array (Illumina Inc, San Diego, CA). The quality control process involved filtering for sex and SNPs mismatches, as well as removal of control and blind duplicate samples. Subsequently, methylation data were normalized across all samples using the R package SeSAMe^44^ to mask 105,454 probes, followed by dye bias correction, and normal-exponential deconvolution using out-of-band probes (Noob) background subtraction^45^. The final methylation values were calculated as the beta-values, defined as the percentage of methylation signal for each individual CpG site. The obtained beta-values underwent further correction for type-2 probe bias using beta mixture quantile normalization (BMIQ) from the WateRmelon package^46^, flowed by ComBat batch correction to account for position on array, slide and plate^47^. Cell type proportions were estimated using a reference-based method based on the genome-wide methylation data.

Methylation risk scores (MRS) of CRP were constructed as weighted sums of each individual’s DNAm values of pre-developed CpG sites using *MethParquet* package (v0.1.0)^48^ in R (v4.3.1)^49^, selected via an Elastic Net regression, associated blood CRP level ^21^, and standardized across the sample by subtracting the mean before dividing it by its standard deviation. Of the 1468 CpGs previously identified for constructing the MRS for CRP^21^, 1041 were available for MRS calculation.

#### CRP association analysis

Association analyses were performed using survey linear regression models (*survey* package v4.2.1) ^50^ in R (v4.3.1)^49^ to account for the HCHS/SOL study design and generate inference applicable to the HCHS/SOL target population. Study design features accounted for stratification, sampling weights, and non-response. Blood CRP level was modeled as a continuous variable in all analyses and log-transformed to account for the right skewed distribution. OSA related traits and cognitive phenotypes mentioned above were modeled as continuous variables, while excessive daytime sleepiness (EDS, ESS >10), insomnia (WHIIRS ≥10), hypertension and diabetes were binary indicators.

The associations of PRS- and MRS-CRP with blood CRP level were first validated, followed by estimating the association between sleep traits and related comorbidities with each CRP measure. The primary model 1 adjusted for age, sex, BMI, study center, Hispanic/Latino background, and first 5 genetic PCs in case of PRS and MRS-CRP. Model 2 was fitted further adjusting for hypertension and diabetes status. Statistical significance was based on multiple comparison adjustment with the Benjamini-Hochberg FDR correction on model 1 (FDR q-value<0.05), grouped by each CRP variables. The primary model 1 adjusted for age, sex, BMI, study center, and first 5 genetic ancestry principal components (PCs) in case of PRS and MRS-CRP. Model 2 was fitted further adjusting for hypertension and diabetes status. Statistical significance was based on multiple comparison adjustment with the Benjamini-Hochberg FDR correction on model 1 (FDR q-value<0.05), grouped by each CRP variables.

### Secondary and sensitivity analyses

We estimated the associations of sleep phenotypes that were found to be associated with MRS-CRP in model 1, with diabetes, hypertension and cognitive scores with and without adjusting for MRS-CRP. In addition, lasso penalized regression was applied to select, from the CRP-associated CpG sites used to construct MRS-CRP, those also important to sleep and health outcomes identified in previous association analyses, adjusting for age, sex, BMI, study center, first 5 genetic PCs and six cell-type proportions in blood estimated based on DNA methylation data. Particularly, AHI was modeled as binary variable based on the threshold of 15 to categorize OSA groups into moderate to severe and mild to normal cases.

We also compared MRS-CRP associations with health outcomes from model 1 with and without adjusting for PRS-CRP. To account for potential U-shaped association of sleep duration with MRS- and blood CRP, we fitted penalized cubic spline models using *mgcv* package (v1.9.1)^51^ in R (v4.3.1)^49^, adjusting for the same covariates as in model 1. The basis dimension for the spline term (k) was selected by minimizing the Akaike Information Criterion (AIC).

Using all HCHS/SOL participants (n= 16,415) and those with genotype data (n= 12,343), we repeated the primary association analysis (model 1) for blood-CRP and PRS-CRP respectively, to increase power for detecting associations. Furthermore, we classified OSA cases (AHI>5) according to the absence or presence of EDS cases and tested the association of OSA cases with and without EDS with PRSs, as described in the previous paper^8^.

Given that DNAm profile derived from whole blood represents an average across all present cell types, and the association of systemic inflammation with central obesity and smoking, we performed sensitivity analyses for MRS-CRP adjusting for cell-type proportions, smoking status and waist-hip ratio, respectively.

## Results

### Demographics and outcome characteristics of study samples

Sample characteristics for MESA are summarized in Table S1. The number of participants in the three CVD risk groups is evenly distributed. There are race/ethnic differences in CRP levels, with the proportion of Chinese individuals decreasing by 18% while the proportion of Black and Hispanic individuals increased by 10% between the low risk and elevated risk groups.

Descriptive statistics of demographic characteristics in the primary analytical sample are provided in Table 1. Overall, women constitute 65.8% of this sample, with higher female percentage, 75.7%, among participants with blood CRP level larger than 3 mg/L. Note that the high proportion of women is due to prioritization of individuals with Mild Cognitive Impairment (MCI) into the SOL-INCA methylation study, combined with higher MCI prevalence in women compared to men. Most OSA related measurements, along with hypertension and diabetes, exhibit greater severity and incidence rate in parallel with higher CRP blood level, whereas sleep duration, insomnia, daytime sleepiness and cognitive scores show little difference across the three risk groups (Table 1).

**Table 1.** Demographic characteristics of HCHS/SOL stratified by cardiovascular risk groups based on C-reactive protein levels (mg/L)

|  | **Overall**<br>(n = 2221) | **Low risk (<1)**<br>(n = 456) | **Borderline (1-3)**<br>(n = 849) | **Elevated risk (>3)**<br>(n = 916) |
| --- | --- | --- | --- | --- |
| Age (Mean (SD)) | 56.66 (8.22) | 56.77 (8.33) | 56.57 (8.10) | 56.69 (8.26) |
| BMI (Mean (SD)) | 30.23 (5.51) | 26.86 (3.79) | 29.38 (4.57) | 32.70 (5.87) |
| Sex (%) - Female | 1435 (58.4) | 227 (44.9) | 520 (53.6) | 688 (69.8) |
| CRP (mg/L) (Mean (SD)) | 4.32 (10.30) | 0.60 (0.21) | 1.87 (0.58) | 8.44 (15.07) |
| **Background (%)** |  |  |  |  |
| Central American | 214 (6.1) | 44 (5.4) | 78 (6.9) | 92 (5.6) |
| Cuban | 380 (28.4) | 70 (25.0) | 126 (25.0) | 184 (33.2) |
| Dominican | 241 (10.2) | 49 (10.6) | 93 (10.1) | 99 (10.0) |
| Mexican | 754 (26.9) | 178 (34.4) | 313 (29.2) | 263 (20.9) |
| Puerto Rican | 432 (20.6) | 77 (17.4) | 156 (19.8) | 199 (23.1) |
| South American | 159 (6.1) | 32 (5.4) | 64 (6.9) | 63 (5.7) |
| **Sleep traits (mean (SD))** |  |  |  |  |
| AHI | 8.16 (12.46) | 6.11 (10.37) | 7.41 (10.80) | 9.94 (14.48) |
| Min SpO2 | 85.75 (6.49) | 86.67 (6.34) | 86.26 (6.14) | 84.78 (6.77) |
| Average SpO2 | 96.29 (0.89) | 96.46 (0.81) | 96.34 (0.80) | 96.15 (1.0) |
| % Time SpO2<90 | 1.03 (2.99) | 0.69 (2.34) | 0.88 (2.67) | 1.36 (3.53) |
| ESS<sup>a)</sup> | 6.07 (4.96) | 5.95 (4.86) | 5.94 (4.94) | 6.25 (5.04) |
| WHIIRS<sup>b)</sup> | 7.96 (5.57) | 7.52 (5.37) | 7.78 (5.42) | 8.35 (5.78) |
| Sleep Duration (Mean (SD)) | 7.80 (1.43) | 7.70 (1.30) | 7.79 (1.36) | 7.85 (1.56) |
| **Comorbidities (%)** |  |  |  |  |
| Diabetes | 678 (30.8) | 101 (22.0) | 235 (28.6) | 342 (37.3) |
| Hypertension | 988 (46.1) | 163 (37.2) | 367 (42.8) | 458 (53.7) |
| **Cognitive traits (Mean (SD))** |  |  |  |  |
| Global Cognitive Function |  |  |  |  |
| Baseline | 0.06 (0.76) | 0.06 (0.76) | 0.07 (0.77) | 0.06 (0.75) |
| Global Cognitive Change | -0.18 (0.56) | -0.14 (0.53) | -0.19 (0.58) | -0.18 (0.57) |

a) ESS: Epworth Sleepiness Scale, b) WHIIRS: Women's Health Initiative Insomnia Rating

### Performance-based selection of PRS for CRP

Table S2 provides the number of SNPs used in different PRSs. The best performing PRS-CRP was selected as the one with the highest CRP blood level variance explained in MESA (Table S3). European ancestry-specific PRS-CSx explained more than 9% of the CRP level. Consequently, the PRS-CSx of European ancestry (PRS-EUR) wa selected for subsequent association analysis in HCHS/SOL dataset. However, previous multi-cohort studies have shown that a weighted sum trained over multiple PRSs yields stronger association with related phenotypes. Also, PRS weighted summation is the intended, developed use of PRS-CSx PRSs. Therefore, in HCHS/SOL, we also constructed a weighted sum of the five ancestry-specific PRS-CSx with adaptive weights (PRS-wsum), obtained using all MESA participants (PRS-wsum-All) and only MESA Hispanic participants (summation weights are provided in Table S4). Because PRS-wsum-all and PRS-wsum-Hispanic had comparable associations with blood CRP level in HCHS/SOL (Figure S1), we focus PRS-wsum-all in subsequent association analyses. Table S5 provides the number of SNPs used for PRS-CSx construction in HCHS/SOL.

Associations between PRS, MRS-CRP and blood CRP level. Both PRS and MRS-CRP are associated with blood CRP level (Figure 2). MRS-CRP showed an almost two-fold higher association than the two PRSs, yielding a coefficient of 1.43 mg/L per one standard deviation increase in MRS-CRP. On the other hand, one standard deviation increase in weighted sum of PRS-CRP amounts to 1.23 mg/L increase in blood CRP level.

**Figure 2.**
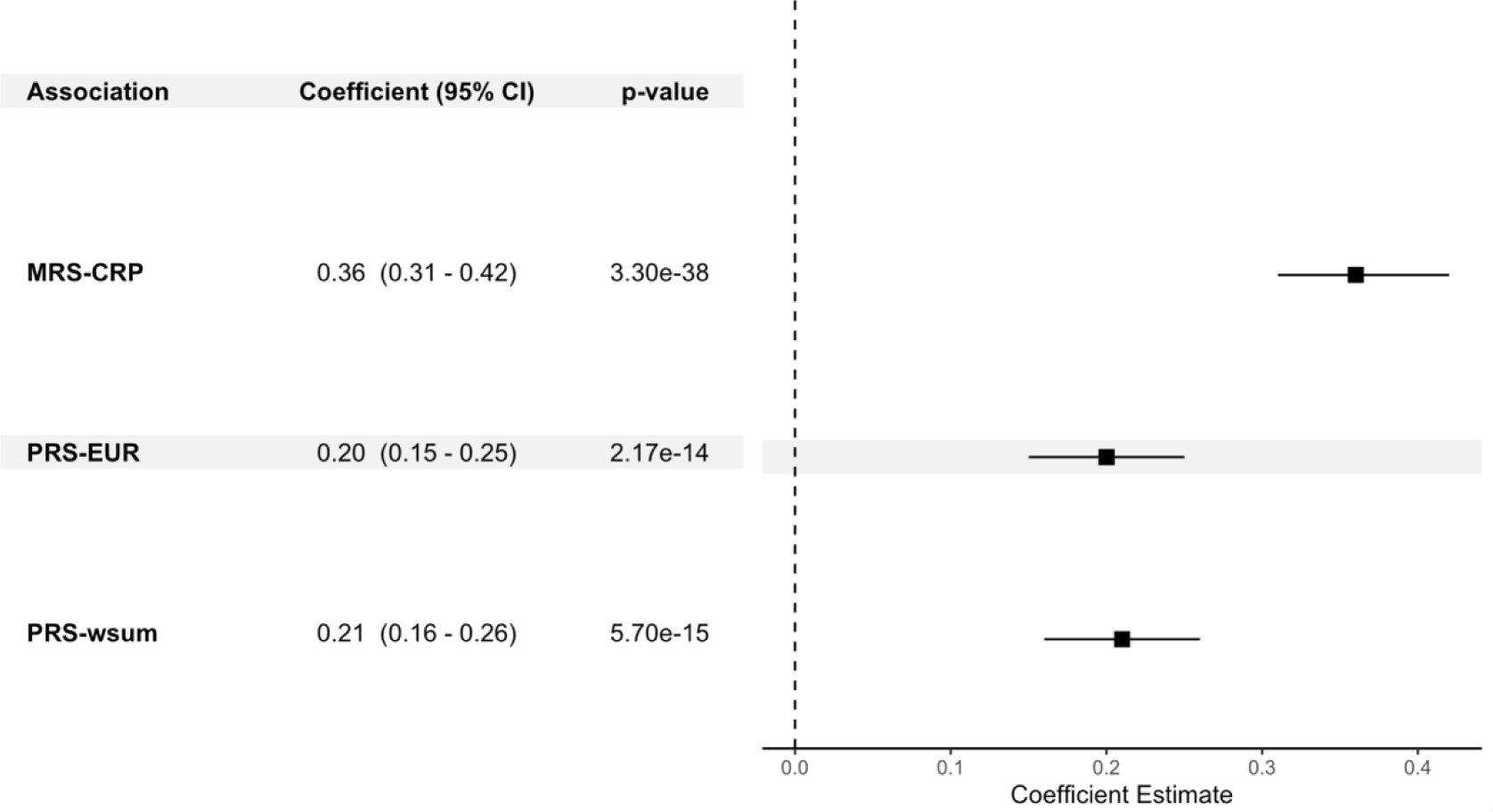
Association of polygenic risk score (PRS) and methylation risk score (MRS) for C-reactive protein (CRP) with blood CRP level.

CRP associations with sleep and other comorbidities. Figure 3 summarizes the association between the CRP measures and phenotypic outcomes with corresponding 95% confidence intervals. Interestingly, MRS-CRP is the only metric showing statistically significant association with AHI (1.11 [0.36, 1.86]), minimum (-0.56 [-1.05, -0.08]) and average (-0.10 [-0.17, -0.04]) SpO2 (Figure 3A), implying that MRS-CRP correlates positively with OSA severity. Although not exhibiting significant associations with OSA traits, the coefficient of blood CRP level displays the same direction as that of MRS-CRP.

**Figure 3.**
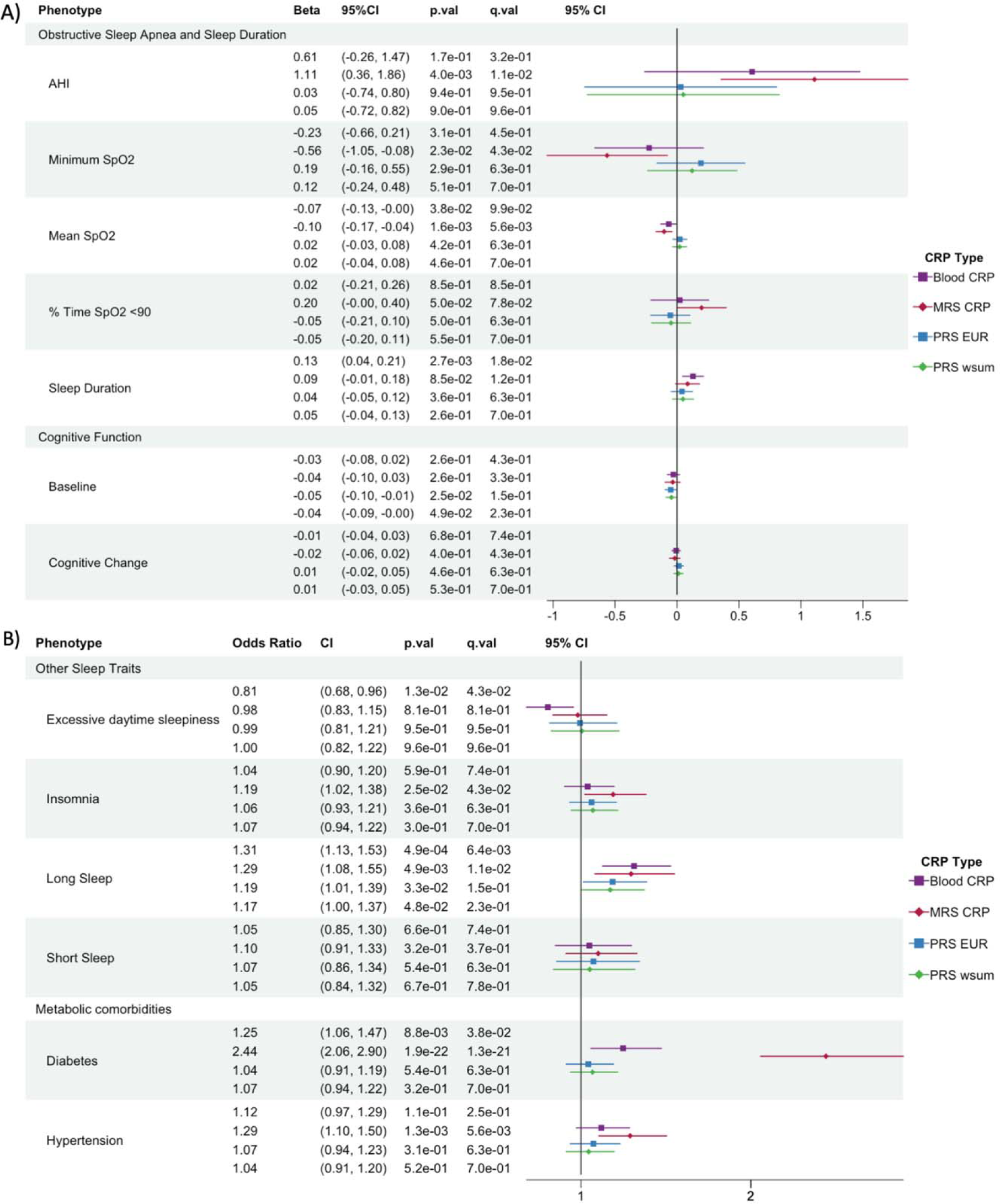
Forest plot showing associations between C-reactive protein (CRP) measures and measured health outcomes. A) obstructive sleep apnea (OSA), sleep duration and cognitive traits; B) odds ratio for binary cardio-metabolic and other sleep outcomes. From left to right model coefficients or odds ratio in case of binary outcomes, 95% confidence interval (95% CI), p value (p.val) and FDR corrected q value (q.val). MRS: methylation risk score; PRS: polygenic risk score

As shown in Figure 3B, MRS-CRP is also associated with insomnia (odds ratio (OR) = 1.19 [1.02, 1.38]) and long sleep duration (OR = 1.29 [1.08, 1.55]), while blood CRP level is associated with EDS (OR = 0.81 [0.68, 0.96]) and overall sleep duration (0.13 [0.04, 0.21]), and long sleep duration (OR = 1.31, [1.13, 1.53]). Meanwhile, short sleep duration is not significantly associated with any of the CRP measures (Figure 3B). On the other hand, PRS-CRPs are not associated with any sleep traits (Figure 3).

Regarding other comorbidities, the relationship between individuals with greater genetic susceptibility to CRP, indicated by higher standardized weighted sum of PRS-CRP, and lower cognitive function at baseline (-0.05 [-0.1, -0.01]) fails to pass the multiple testing correction (Figure 3A). Surprisingly, MRS-CRP remains the only CRP measure to have strong associations (q-value<0.001) with both cardio-metabolic diseases, diabetes (OR = 2.44, [2.06, 2.90]) and hypertension (OR = 1.29, [1.10, 1.50]) (Figure 3B).

The effect of MRS-CRP on the health outcomes decreased slightly in model 2 (Figure S2). More particularly, its associations with OSA and diabetes remain statistically significant after adjusting for cardio-metabolic diseases, except for minimum SpO2, insomnia, long sleep duration and hypertension (Figure S2).

### Secondary analyses

#### Association between sleep and related comorbidities

AHI and minimum SpO2 were associated with diabetes, cognitive function and hypertension, and average SpO2 was associated with the two cardio-metabolic diseases (Figure S3A). Notably, cognitive function score at baseline was positively associated with OSA severity, yet inversely associated with insomnia and long sleep duration (Figure S3A). Adjusting for MRS-CRP reduced the effect size of the associations with the two cardio-metabolic disorders (Figure S3B), suggesting that MRS-CRP captures the inflammatory component shared by sleep phenotypes and these comorbidities. Interestingly, the strongest change was observed for diabetes, whereas adjusting for MRS-CRP had a much smaller impact on associations with hypertension and cognitive scores (Figure S3). Adjusting for PRS-CRP essentially had no effect on the associations of MRS-CRP with OSA phenotypes, long sleep and metabolic comorbidities (Figure S4).

#### Cubic spline model for sleep d uration

Our cubic spine model results supported the previously reported U-shaped (or J-shaped) association between sleep duration and CRP markers^52,53^, where optimal sleep time (around 7 hours) corresponds to lowest level of both MRS- and blood CRP level (Figure S5). The more robust relationship between long sleep and CRP markers (Figure S5) agrees with a previous study supporting the J-shaped association^52^. The protective effect of short sleep on blood CRP persisted in analyses that utilized data from the larger sample, while the rate of increase in blood CRP level associated with longer sleep becomes less pronounced (Figure S5).

#### Association analysis using all HCHS/SOL participants

Interestingly, associations of blood CRP level with AHI (0.41 [0.16, 0.66]) and cognitive function at baseline (-0.04 [-0.06, -0.01]) become statistically significant after increasing the sample size to all HCHS/SOL participants, yet its association with EDS become statistically insignificant (Figure S6). When it comes to PRS-CRPs, their association p-values remain unchanged across these health outcomes (Figure S6). Table S6 provides the results of association of PRS-CRPs with OSA risk, with and without EDS, in comparison with no OSA. In contrast to the previous study^8^, our computed PRS-CRP was negatively associated with OSA with EDS (Table S6).

#### Lasso penalized regression

Out of the list of the 1,041 CpG sites used in the construction of the MRS-CRP score, lasso penalized regression identified 66 and 12 CpG sites as pertinent to AHI and minimum SpO2, respectively, yet 0 CpG sites for average SpO2 and insomnia (Table S7-S8). In terms of diabetes and hypertension, 55 and 6 CRP associated CpG sites were selected by lasso penalized regression (results not shown). Among them, 6 CpG sites are shared between OSA related measures, and 5 shared across OSA and one of the two metabolic disorders (Table 2).

**Table 2.** CpGs selected by Lasso regression across obstructive sleep apnea and metabolic comorbidities.

| CpG | Chromosome | Gene Name | Phenotypes |
| --- | --- | --- | --- |
| cg00572560 | 10 |  | AHI, Diabetes |
| cg00574958 | 11 | CPT1A | AHI, Min SpO2, Diabetes |
| cg03246954 | 19 | MKNK2 | AHI, Min SpO2 |
| cg06690548 | 4 | SLC7A11 | AHI, Hypertension |
| cg14476101 | 1 | PHGDH | AHI, Min SpO2, Hypertension |
| cg14656297 | 9 | FXN | AHI, Min SpO2 |
| cg19693031 | 1 | TXNIP | AHI, Diabetes, Hypertension |
| cg23281327 | 10 |  | AHI, Min SpO2 |
| cg23440058 | 3 | KALRN | AHI, Min SpO2 |

#### Sensitivity analysis

Adjusting for cell-type proportions, smoking status and waist-hip ratio yielded comparable results in association significance level to that of primary analysis for MRS-CRP (Figure S7). Surprisingly, using waist-hip ratio as parameter for obesity instead of BMI augmented considerably the association between MRS-CRP and these health outcomes except for diabetes (Figure S7).

## Discussion

MRS-CRP stands out as the only CRP measure associated with insomnia and OSA severity markers including AHI, minimum and average SpO2. Blood CRP level, on the other hand, is only associated with AHI after increasing the sample pool to all HCHS/SOL participants and not associated with average SpO2 in either participants with OSA^54^ or the general Hispanic population. That said, blood CRP is likely a less specific and accurate marker for OSA compared to MRS-CRP. Our results supported the superior performance of MRS-CRP to circulating CRP in measuring chronic inflammation and comorbidities^18–21^.

Several factors could impact the performance of circulating CRP. First of all, fluctuations in circulating CRP levels are highly susceptible to circadian misalignment, acute medical condition and genetic predisposition^55,56^. Furthermore, CRP molecules exist in two structural forms, pentameric and monomeric^57^. The former is synthesized in the liver and regulated by proinflammatory cytokines. While the latter is dissociated from the pentameric CRP at local inflammation sites such as atherosclerotic plaque, further escalating inflammatory activities^58^. However, current high sensitivity assays measure only the pentameric form^57^, thus limiting the representation of overall inflammatory status. In contrast, MRS-CRP, derived from a mixture of cellular populations in the blood, reflects epigenetic changes resulting from cellular responses to inflammation. This makes MRS-CRP a promising marker for stratifying population risk across numerous outcomes, including chronic inflammation, sleep and metabolic health, thereby enhancing accuracy and clinical inference.

CRP has emerged as a useful clinical marker for OSA and cardiovascular comorbidities, due to its independent association with apnea severity and hypertension^59^. Given that OSA is recognized as an antecedent risk factor for secondary hypertension^60^, the modest change in association between OSA traits and hypertension after adjusting for MRS-CRP suggests a relatively small contribution of inflammation to the elevated blood pressure. Nevertheless, the association between sleep outcomes and diabetes were greatly attenuated and even no longer statistically significant after adjusting for MRS-CRP, which is in alignment with the mitigating effect of CRP over the association of OSA severity and incident diabetes^61^. These findings indicate that systemic inflammation may function as a mediator or a common factor underlying the relationship between sleep phenotypes and diabetes.

Both MRS- and blood CRP markers manifest a J-shaped association with sleep duration. Previous research has linked diabetes, hypertension, depression and low-grade inflammation^62^ to prolonged sleep time, which is generally considered as an indicator of poor health condition^63^. Congruent with our findings, the relationship between short sleep duration and CRP levels is no longer statistically significant after adjusting for confounding variables^52,53^, probably due to the strong associations of sex, race/ethnicity, and age with both extreme sleep durations and blood CRP level^52^. Interestingly, the slightly protective effect of short sleep on CRP levels in Hispanic (Figure S5) and Asian population^52^ warrants future studies to replicate these results using larger sample sizes among different races/ethnic groups, and to investigate how inflammation interplays with sleep duration and cardiovascular comorbidities.

The non-significant associations observed for PRS-CRP with sleep and health outcomes indicate a weak relationship between genetic component of CRP with related comorbidities. Circulating CRP level is thus likely the consequence of these health conditions^9,64^, not the other way around. One possible reason for the inconsistent association results of PRS-CRP with OSA cases with EDS from the previous paper^8^ is the distinct assessment criteria for OSA and EDS employed by the two studies, aside from intrinsic differences across sampled populations. In the study conducted by Huang and colleagues ^8^, OSA cases were based on clinical diagnosis and their EDS assessment showed only a moderate correlation with ESS. Nevertheless, PRS-CRP showed a significant association with blood pressure, albeit not with diabetes, in European population^65^. These conflicting results emphasize the need for more research to develop PRS-CRPs and estimate their associations across populations of different ancestries and extra caution in its application for clinical assessment and inference.

Given that sleep questionnaires, OSA assessments, blood sampling for CRP DNAm assays were all conducted during the baseline visit, we were limited to performing association analysis. Another limitation of current study stems from the potential bias in self-reported sleep measurements, which often fall short of capturing context-specific sleep patterns influenced by environmental and lifestyle factors. Overall, it remains to be determined whether the inflammation-associated DNAm causes or results from sleep and metabolic disorders, or if a third scenario exists where both are affected by a shared underlying causal factor. In light of the established associations, the underlying epigenetic mechanisms behind the observed inflammatory processes and how they relate to sleep and its co-morbidities still need to be elucidated.

To address this question, we applied lasso regression to select key CRP-associated CpGs related to health outcomes. Consequently, 6 CpGs were found to be shared between OSA traits and metabolic disorders. Specifically, cg19693031 methylation in the thioredoxin interacting protein (TXNIP) gene is inversely associated with HbA1c^66^, fasting blood glucose^67^, triglycerides levels^66^ and blood pressure^68^. Various studies confirmed its strong association with incident and prevalent type 2 diabetes^69–71^. Hypomethylation of cg19693031 enhances TXNIP expression, which in turn promotes inflammation and activates monocytes towards an inflammatory state ^71,72^. The negative coefficient of this locus obtained by lasso regression supports its protective role against the disease onset (Table S7-S9). This suggests a possible connection between OSA and diabetes through hyperglycemia-induced inflammation^73^.

Hypermethylation at cg00574958 of carnitine palmitoyltransferase-1A (CPT1A) also showed a protective association with AHI, minimum SpO2, and diabetes (Table S7-S9), consistent with previous studies of diabetes^74,75^. CPT1A mediates energy homeostasis by regulating the rate of mitochondrial fatty acid oxidation^76^. A multi-cohort study reported associations between carbohydrate and fat intake with changes at CPT1A-cg00574958 locus, with implication in mediating risks of metabolic traits including BMI, diabetes, hypertension, as well as triglyceride and glucose levels^74^.

Solute carrier family 7 member 11 (SLC7A11)-cg06690548 and phosphoglycerate dehydrogenase (PHGDH)-cg14476101 have been frequently reported together as showing protective association against blood pressure^77,78^. Such protective effect was also observed in HCHS/SOL, with hypermethylation at both sites associated with improved AHI and SpO2 levels, as well as lower hypertension risk (Table S7-S9). PHGDH-cg14476101 is associated with triglyceride level^79^ and urinary albumin-to-creatinine ratio^78^ by regulating serine biosynthesis and lipid homeostasis^80^. DNAm change in cg06690548 affects the expression of SLC7A11 to modulate metabolic reprogramming and the defense mechanism against oxidative stress^77,81^.

Overall, these lasso-selected CpG sites reiterate the important role of DNAm in the pathogenesis underlying complex diseases, and may serve as potential biomarkers for inflammatory stress associated with metabolic syndrome and OSA traits, bridged by fat, glucose and energy homeostasis. The large effect sizes of some CpGs for AHI and minimum SpO2, such as MKNK2-cg03246954, PFDN2-cg00816397, ATF2-cg02622866 and ARSA-cg07453718, indicate strong associations with OSA, and potential applicability as disease biomarkers (Table S7-S8). Future research could benefit from confirming causality of these associations as well as their directionality, and expanding to populations with other ancestries to test efficacy of the epigenetic predictor of CRP.

## Conclusion

A CRP measure based on DNA methylation status at select CpG sites outperforms the genetic and blood measure of circulating CRP in association with several OSA-related sleep traits and metabolic comorbidities commonly linked to chronic inflammation. However, associations with other sleep and cognitive measures were mixed. Our results also suggest a probable shared mechanism between sleep phenotypes and metabolic syndrome, related to systemic inflammation and modeled by MRS-CRP, particularly diabetes. Consequently, the incorporation of MRS-CRP, versus circulating CRP, in clinical assessment has the potential to improve population risk stratification for adverse health outcomes

## Data Availability

HCHS/SOL data are available through application to the data base of genotypes and phenotypes (dbGaP) accession phs000810, or via data use agreement with the HCHS/SOL Data Coordinating Center (DCC) at the University of North Carolina at Chapel Hill, see collaborators website: https://sites.cscc.unc.edu/hchs/.
TOPMed-MESA freeze 10 WGS data are available by application to dbGaP according to the study specific accession: MESA: "phs001211". MESA phenotype data are available from dbGaP according to the MESA accession: MESA: "phs000209".
Summary statistics from GWAS of CRP were downloaded from the PAN-UKBB project (https://pan.ukbb.broadinstitute.org/; phenocode 30710) and BioBank Japan PheWeb at https://pheweb.jp/pheno/CRP.
PRS-CRP alleles and weights are provided at the Github repository: https://github.com/ZWangTen/CRP_markers_association.

## Data Availability

HCHS/SOL data are available through application to the data base of genotypes and phenotypes (dbGaP) accession phs000810, or via data use agreement with the HCHS/SOL Data Coordinating Center (DCC) at the University of North Carolina at Chapel Hill, see collaborators website: https://sites.cscc.unc.edu/hchs/.

TOPMed-MESA freeze 10 WGS data are available by application to dbGaP according to the study specific accession: MESA: “phs001211”. MESA phenotype data are available from dbGaP according to the MESA accession: MESA: “phs000209”.

Summary statistics from GWAS of CRP were downloaded from the PAN-UKBB project (https://pan.ukbb.broadinstitute.org/; phenocode 30710) and BioBank Japan PheWeb at https://pheweb.jp/pheno/CRP.

PRS-CRP alleles and weights will be provided at the Github repository: https://github.com/ZWangTen/CRP_markers_association.

## Acknowledgement

The authors thank the staff and participants of MESA and HCHS/SOL for their important contributions. HCHS/SOL investigators website - http://www.cscc.unc.edu/hchs/. The work was supported by National Heart Lung and Blood Institute (NHLBI) grants R01HL161012 to T.S., National Institute on Aging (NIA) grants R01AG080598 to T.S., RF1AG061022 to MF and HMG, and R01AG048642, R56AG048642 and R01AG075758 to HMG. The Hispanic Community Health Study/Study of Latinos is a collaborative study supported by contracts from the National Heart, Lung, and Blood Institute (NHLBI) to the University of North Carolina (HHSN268201300001I / N01-HC-65233), University of Miami (HHSN268201300004I / N01-HC-65234), Albert Einstein College of Medicine (HHSN268201300002I / N01-HC-65235), University of Illinois at Chicago (HHSN268201300003I / N01-HC-65236 Northwestern Univ), and San Diego State University (HHSN268201300005I / N01-HC-65237). The following Institutes/Centers/Offices have contributed to the HCHS/SOL through a transfer of funds to the NHLBI: National Institute on Minority Health and Health Disparities, National Institute on Deafness and Other Communication Disorders, National Institute of Dental and Craniofacial Research, National Institute of Diabetes and Digestive and Kidney Diseases, National Institute of Neurological Disorders and Stroke, NIH Institution-Office of Dietary Supplements.

MESA and the MESA SHARe project are conducted and supported by the National Heart, Lung, and Blood Institute (NHLBI) in collaboration with MESA investigators. Support for MESA is provided by contracts HHSN268201500003I, N01-HC-95159, N01-HC-95160, N01-HC-95161, N01-HC-95162, N01-HC-95163, N01-HC-95164, N01-HC-95165, N01-HC-95166, N01-HC-95167, N01-HC-95168, N01-HC-95169, UL1-TR-000040, UL1-TR-001079, UL1-TR-001420. MESA Family is conducted and supported by the National Heart, Lung, and Blood Institute (NHLBI) in collaboration with MESA investigators. Support is provided by grants and contracts R01HL071051, R01HL071205, R01HL071250, R01HL071251, R01HL071258, R01HL071259, and by the National Center for Research Resources, Grant UL1RR033176. The provision of genotyping data was supported in part by the National Center for Advancing Translational Sciences, CTSI grant UL1TR001881, and the National Institute of Diabetes and Digestive and Kidney Disease Diabetes Research Center (DRC) grant DK063491 to the Southern California Diabetes Endocrinology Research Center. Molecular data for the Trans-Omics in Precision Medicine (TOPMed) program was supported by the National Heart, Lung and Blood Institute (NHLBI). Genome sequencing for “NHLBI TOPMed: Multi-Ethnic Study of Atherosclerosis” (phs001416.v2.p1) was performed at Broad Institute Genomics Platform (HHSN268201500014C, 3U54HG003067-13S1. Core support including centralized genomic read mapping and genotype calling, along with variant quality metrics and filtering were provided by the TOPMed Informatics Research Center (3R01HL-117626-02S1; contract HHSN268201800002I). Core support including phenotype harmonization, data management, sample-identity QC, and general program coordination were provided by the TOPMed Data Coordinating Center (R01HL-120393; U01HL-120393; contract HHSN268201800001I). We gratefully acknowledge the studies and participants who provided biological samples and data for TOPMed.

## Ethics Statement

HCHS/SOL: This study was approved by the institutional review boards (IRBs) at each field center, where all participants gave written informed consent, and by the Non-Biomedical IRB at the University of North Carolina at Chapel Hill, to the HCHS/SOL Data Coordinating Center. All IRBs approving the study are: Non-Biomedical IRB at the University of North Carolina at Chapel Hill. Chapel Hill, NC; Einstein IRB at the Albert Einstein College of Medicine of Yeshiva University. Bronx, NY; IRB at Office for the Protection of Research Subjects (OPRS), University of Illinois at Chicago. Chicago, IL; Human Subject Research Office, University of Miami. Miami, FL; Institutional Review Board of San Diego State University. San Diego, CA.

MESA: All MESA participants provided written informed consent, and the study was approved by the Institutional Review Boards at The Lundquist Institute (formerly Los Angeles BioMedical Research Institute) at Harbor-UCLA Medical Center, University of Washington, Wake Forest School of Medicine, Northwestern University, University of Minnesota, Columbia University, and Johns Hopkins University.

This work was approved by the Beth Israel Deaconess Medical Center Committee on Clinical Investigators (protocol #2023P000538).

