## Supplementary File for "Analysis of C-reactive protein omics-measures associates methylation risk score with sleep health and related health outcomes"

Supplementary information

### Supplementary Tables

#### Table S1. Demographic characteristics of MESA

|  | Overall  (n = 6411) | Low risk (<1)  (n = 1926) | Borderline (1-3)  (n = 2184) | Elevated risk (>3)  (n = 2301) |
| --- | --- | --- | --- | --- |
| Age (Mean (SD)) | 62.2 (10.2) | 61.5 (10.7) | 62.9 (10.2) | 62.3 (9.8) |
| BMI (Mean (SD)) | 28.3 (5.5) | 25.6 (4.2) | 28.0 (4.6) | 30.8 (6.0) |
| CRP (mg/L) (Mean (SD)) | 3.8 (5.9) | 0.6 (0.2) | 1.8 (0.6) | 8.3 (8.0) |
| Gender (%) - Female | 52.4 | 42 | 48 | 65.1 |
| Race/ethnicity (%) | | | | |
| White | 39.3 | 41.2 | 39.9 | 37.1 |
| Black | 26 | 20.2 | 24.6 | 32.3 |
| Chinese | 12.1 | 22.2 | 11.5 | 4.2 |
| Hispanic/Latino | 22.6 | 16.3 | 24 | 26.4 |

| Table S2. SNP counts in polygenic risk score (PRS) construction for MESA | | | |
| --- | --- | --- | --- |
| Threshold/  Type | R2 | Distance  (Kb) | # SNP |
| BBJ+UKBB | | | |
| 5.00E-08 | 0.1 | 1000 | 1162 |
| 1.00E-07 |  |  | 1249 |
| 1.00E-06 |  |  | 1760 |
| 1.00E-05 |  |  | 2956 |
| 1.00E-04 |  |  | 6186 |
| 1.00E-03 |  |  | 17126 |
| 1.00E-02 |  |  | 57357 |
| 1.00E-01 |  |  | 197806 |
| PRS_CSx | | | |
| AFR | / | / | 1165122 |
| EUR |  |  | 1055663 |
| AMR |  |  | 1119543 |
| PRS_Huang et.al | | | |
| Weighted sum | / | / | 48 |

| Table S3. Association analysis for polygenic risk score (PRS)-CRP and blood CRP level in MESA | | | | |
| --- | --- | --- | --- | --- |
| PRS  Threshold/type | **PRS**  **estimate** | **SE** | **p value** | **Variance**  **Explained (%)** |
| BBJ + UKBB, with clumping | | | | |
| 5.00E-08 | 0.24 | 0.01 | 2.2E-73 | 5.38 |
| 1.00E-07 | 0.24 | 0.01 | 1.3E-70 | 5.63 |
| 1.00E-06 | 0.21 | 0.01 | 2.3E-50 | 4.48 |
| 1.00E-05 | 0.22 | 0.02 | 9.3E-23 | 3.64 |
| 1.00E-04 | 0.19 | 0.03 | 1.2E-09 | 2.65 |
| 1.00E-03 | 0.19 | 0.05 | 1.0E-04 | 2.32 |
| 1.00E-02 | 0.21 | 0.07 | 1.7E-03 | 2.2 |
| 1.00E-01 | 0.21 | 0.08 | 1.0E-02 | 2.16 |
| BBJ + UKBB, without clumping | | | | |
| AFR | 0.03 | 0.01 | 1.8E-02 | 0.004 |
| EUR | 0.31 | 0.02 | 8.3E-83 | 9.33 |
| EAS | 0.1 | 0.01 | 3.54E-15 | 1.73 |
| SAS | 0.09 | 0.02 | 1.61E-08 | 0.7 |
| AMR | 0.002 | 0.02 | 8.7E-01 | 0.21 |
| 48 SNPs from Huang et al. | | | | |
| PRS_Huang | 0.25 | 0.01 | 6.2E-77 | 6.0 |

#### Table S4. Polygenic risk score (PRS) effect size obtained using MESA cohort

| PRS_CSx | Estimate | Std. Error | p value | Group |
| --- | --- | --- | --- | --- |
| AFR | 0.031 | 0.014 | 0.03 | All MESA |
| EUR | 0.288 | 0.016 | 5.11E-69 | All MESA |
| AMR | 0.001 | 0.015 | 0.93 | All MESA |
| EAS | 0.05 | 0.013 | 0.00012 | All MESA |
| SAS | 0.023 | 0.015 | 0.14 | All MESA |
| AFR | 0.042 | 0.028 | 0.13 | MESA Hispanic |
| EUR | 0.267 | 0.032 | 4.24E-16 | MESA Hispanic |
| AMR | -0.007 | 0.029 | 0.79 | MESA Hispanic |
| EAS | 0.078 | 0.026 | 0.003 | MESA Hispanic |
| SAS | 0.061 | 0.029 | 0.038 | MESA Hispanic |

#### Table S5. SNP counts in polygenic risk score (PRS) construction for HCHS/SOL

| Threshold/  Type | R2 | Distance  (Kb) | # SNP |
| --- | --- | --- | --- |
| PRS_CSx | | | |
| AFR | / | / | 333979 |
| EAS |  |  | 302719 |
| EUR |  |  | 320917 |
| AMR |  |  | 330219 |
| SAS |  |  | 322661 |

#### Table S6. Association analysis for obstructive sleep apnea (OSA) cases with and without excessive daytime sleepiness (EDS) in HCHS/SOL

| Comparison | PRS type | Odds ratio | p value | 95% CI | Sample size |
| --- | --- | --- | --- | --- | --- |
| OSA with EDS vs no OSA | PRS-ty | 1.04 | 0.61 | (0.89, 1.21) | 8234 |
| OSA without EDS vs no OSA | PRS-ty | 0.98 | 0.55 | (0.9, 1.06) | 10274 |
| All OSA vs no OSA | PRS-ty | 0.99 | 0.86 | (0.92, 1.07) | 10956 |
| OSA with EDS vs no OSA | PRS-EUR | 0.88 | 0.05 | (0.78, 1) | 8234 |
| OSA without EDS vs no OSA | PRS-EUR | 0.96 | 0.3 | (0.89, 1.04) | 10274 |
| All OSA vs no OSA | PRS-EUR | 0.94 | 0.11 | (0.88, 1.01) | 10956 |
| OSA with EDS vs no OSA | PRS-wsum | 0.85 | 0.02 | (0.75, 0.97) | 8234 |
| OSA without EDS vs no OSA | PRS-wsum | 0.96 | 0.25 | (0.89, 1.03) | 10274 |
| All OSA vs no OSA | PRS-wsum | 0.94 | 0.07 | (0.88, 1) | 10956 |

OSA: obstructive sleep apnea. EDS: excessive daytime sleepiness. PRS-ty: Huang et al. PRS. PRS-EUR: PRS-CSx of European ancestry. PRS-wsum: PRS-CSx with adaptive weights obtained using all MESA participants

#### Table S7. Key CpGs selected by lasso penalized regression for apnea hypopnea index

| CpG | Coefficient | CHR | Gene Name | Overlap |
| --- | --- | --- | --- | --- |
| cg00574958 | -0.39 | 11 | CPT1A | Min SpO2, Diabetes |
| cg14476101 | -0.67 | 1 | PHGDH | Min SpO2, Hypertension |
| cg03246954 | 84.11 | 19 | MKNK2 | Min SpO2 |
| cg14656297 | -8.73 | 9 | FXN | Min SpO2 |
| cg23281327 | -2.98 | 10 |  | Min SpO2 |
| cg23440058 | -1.16 | 3 | KALRN | Min SpO2 |
| cg19693031 | -0.02 | 1 | TXNIP | Diabetes, Hypertension |
| cg00572560 | -1.87 | 10 |  | Diabetes |
| cg06690548 | -0.43 | 4 | SLC7A11 | Hypertension |
| cg00241998 | 0.62 | 4 | N4BP2 |  |
| cg00514616 | 2.63 | 7 |  |  |
| cg00607919 | 2.52 | 2 | CCDC138 |  |
| cg01067983 | -0.48 | 6 | SERPINB6 |  |
| cg01112249 | -7.51 | 10 |  |  |
| cg01526748 | 0.03 | 3 | FGF12 |  |
| cg02622866 | 44.49 | 2 | ATF2 |  |
| cg02761715 | -1.12 | 12 | DUSP16 |  |
| cg03292675 | 0.68 | 8 | EPB49 |  |
| cg05132118 | 22.2 | 13 |  |  |
| cg05828624 | -0.43 | 2 | REG1A |  |
| cg06076692 | 4.39 | 6 | ATXN1 |  |
| cg07001630 | 32.1 | 2 | TMEM214 |  |
| cg07453718 | 52.4 | 22 | ARSA |  |
| cg07794010 | -0.53 | 1 | SYT6 |  |
| cg07817279 | 7.87 | 17 | SFRS1 |  |
| cg07884487 | 14.67 | 3 | MITF |  |
| cg07894567 | -12.88 | 6 |  |  |
| cg08818130 | 3.86 | 3 | ZNF654 |  |
| cg09048665 | -10.84 | 16 | WDR90 |  |
| cg10090326 | -1.89 | 14 | WDR25 |  |
| cg10322504 | -4.57 | 20 |  |  |
| cg10365984 | 0.21 | 6 | BACH2 |  |
| cg10370591 | 1.77 | 2 | TPO |  |
| cg10726559 | -3.04 | 14 | MIR127 |  |
| cg10759591 | 0.08 | 17 | HRNBP3 |  |
| cg12008047 | 1.37 | 19 | GDF15 |  |
| cg12090885 | 1.23 | 3 | GSK3B |  |
| cg12499235 | -0.25 | 11 | ASCL2 |  |
| cg13630239 | 1.2 | 10 | RRP12 |  |
| cg13902024 | -0.03 | 7 | PLXNA4 |  |
| cg14037440 | -0.91 | 9 | FREM1 |  |
| cg14094164 | 1.01 | 13 |  |  |
| cg14532484 | 0.31 | 9 | PAEP |  |
| cg14866339 | -6.71 | 14 | MIR379 |  |
| cg14881305 | -0.82 | 14 |  |  |
| cg15479387 | 0.18 | 6 | LOC285830 |  |
| cg15802555 | -7.12 | 17 | MPP2 |  |
| cg16398761 | -1.59 | 14 | C14orf43 |  |
| cg17087356 | 1.63 | 3 | FHIT |  |
| cg17501395 | 1.21 | 6 | ZC3H12D |  |
| cg17780956 | 2.89 | 4 | MAP9 |  |
| cg18262615 | -15.02 | 3 | KLHL6 |  |
| cg19049696 | -0.19 | 7 | PKD1L1 |  |
| cg19103704 | 0.49 | 19 | FCGBP |  |
| cg19303434 | 9.93 | 4 | SLC30A9 |  |
| cg19756663 | 4.59 | 5 |  |  |
| cg19774973 | -0.49 | 8 |  |  |
| cg20644120 | 15.49 | 12 | SPSB2 |  |
| cg20820543 | -14.21 | 6 | MYB |  |
| cg21877216 | 2.6 | 6 | GNL1 |  |
| cg22980156 | -0.49 | 17 | CDRT4 |  |
| cg25285658 | 0.3 | 4 | MRFAP1L1 |  |
| cg26104690 | -6.04 | 7 | PRKAR2B |  |
| cg26800884 | -1.85 | 4 | KLB |  |
| cg27255239 | 7.84 | 2 | RFX8 |  |
| cg27262415 | 2.82 | 11 | PTPRCAP |  |

#### Table S8. Key CpGs selected by Lasso penalized regression for minimum oxygen saturation

| CpG | Coefficient | CHR | Gene Name | Overlap |
| --- | --- | --- | --- | --- |
| cg00574958 | 0.08 | 11 | CPT1A | AHI, Diabetes |
| cg14476101 | 0 | 1 | PHGDH | AHI, Hypertension |
| cg03246954 | -0.22 | 19 | MKNK2 | AHI |
| cg14656297 | 0.08 | 9 | FXN | AHI |
| cg23281327 | 0.15 | 10 |  | AHI |
| cg23440058 | 0.05 | 3 | KALRN | AHI |
| cg00816397 | -0.76 | 1 | PFDN2 |  |
| cg05802514 | 0.05 | 4 |  |  |
| cg09456254 | 0.01 | 21 | C21orf130 |  |
| cg09728637 | 0.04 | 18 | TYMS |  |
| cg12450708 | 0.02 | 10 |  |  |
| cg24083756 | -0.19 | 21 | MRPS6 |  |

#### Table S9. Shared CpGs selected by lasso penalized regression for diabetes and hypertension

| CpG | Trait | Coefficient | CHR | Gene Name | Overlap |
| --- | --- | --- | --- | --- | --- |
| cg00574958 | Diabetes | -13.54 | 11 | CPT1A | AHI, Min SpO2 |
| cg19693031 | Diabetes | -9.79 | 1 | TXNIP | AHI, Hypertension |
| cg00572560 | Diabetes | 0.39 | 10 |  | AHI |
| cg14476101 | Hypertension | -0.03 | 1 | PHGDH | AHI, Min SpO2 |
| cg19693031 | Hypertension | -0.11 | 1 | TXNIP | AHI, Diabetes |
| cg06690548 | Hypertension | -0.26 | 4 | SLC7A11 | AHI |

### Supplementary Figures

#### Figure S1. Association of polygenic risk score (PRS) and methylation risk score (MRS) for C-reactive protein (CRP) with blood CRP level.

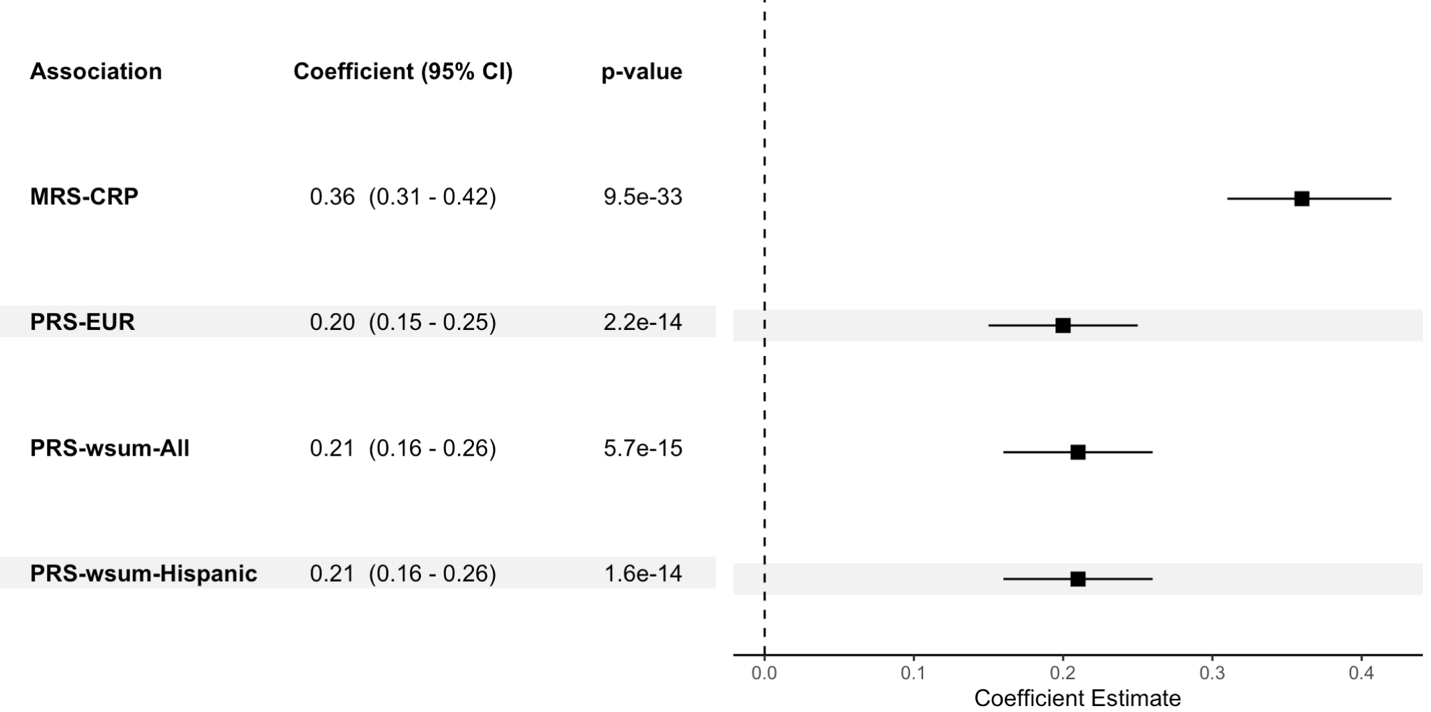

EUR: European ancestry; PRS-wsum-All: PRS calculated as weighted sum of PRSs-CSx with weights obtained using all MESA participants; PRS-wsum-Hispanic: PRS calculated as weighted sum of PRSs-CSx with weights obtained using all Hispanic participants

#### Figure S2. Forest plot comparing effect size of the methylation risk score for C-reactive protein (MRS-CRP) associations obtained using model 1 and model 2 in HCHS/SOL.

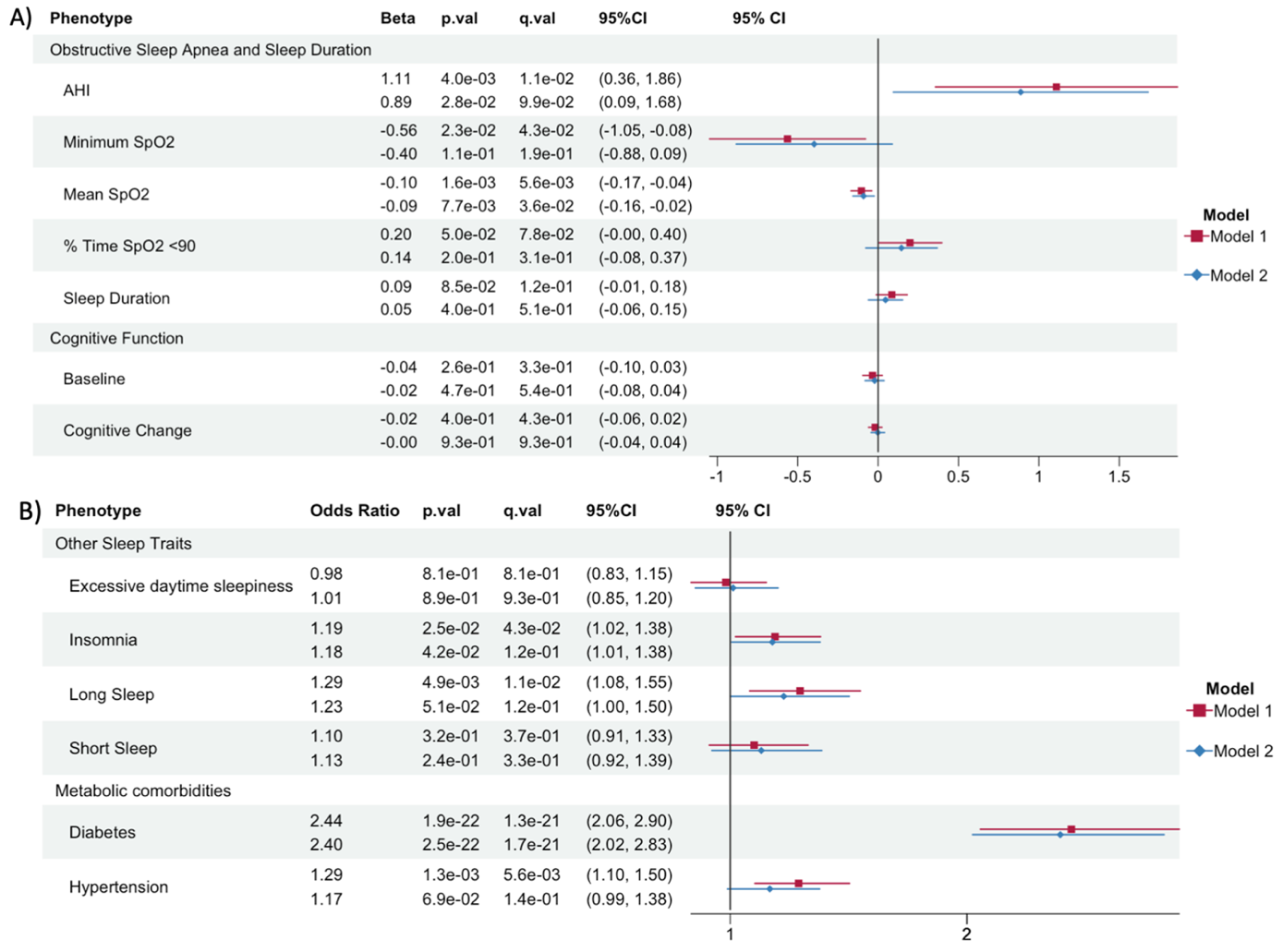

A) obstructive sleep apnea (OSA), sleep duration and cognitive traits; B) odds ratio for binary cardio-metabolic and other sleep outcomes. From left to right model coefficients or odds ratio in case of binary outcomes, p value (p.val), FDR corrected q value (q.val) and 95% confidence interval (95% CI). AHI: apnea hypopnea index; SpO2: oxygen saturation; Cognitive function: change in cognitive function score between baseline and SOL-INCA visit (Change); cognitive function score at baseline (Baseline).

#### Figure S3. Forest plot showing associations of obstructive sleep apnea associated traits with diabetes, hypertension, and cognitive scores with and without adjusting for methylation risk score for C-reactive protein (MRS-CRP) in HCHS/SOL.

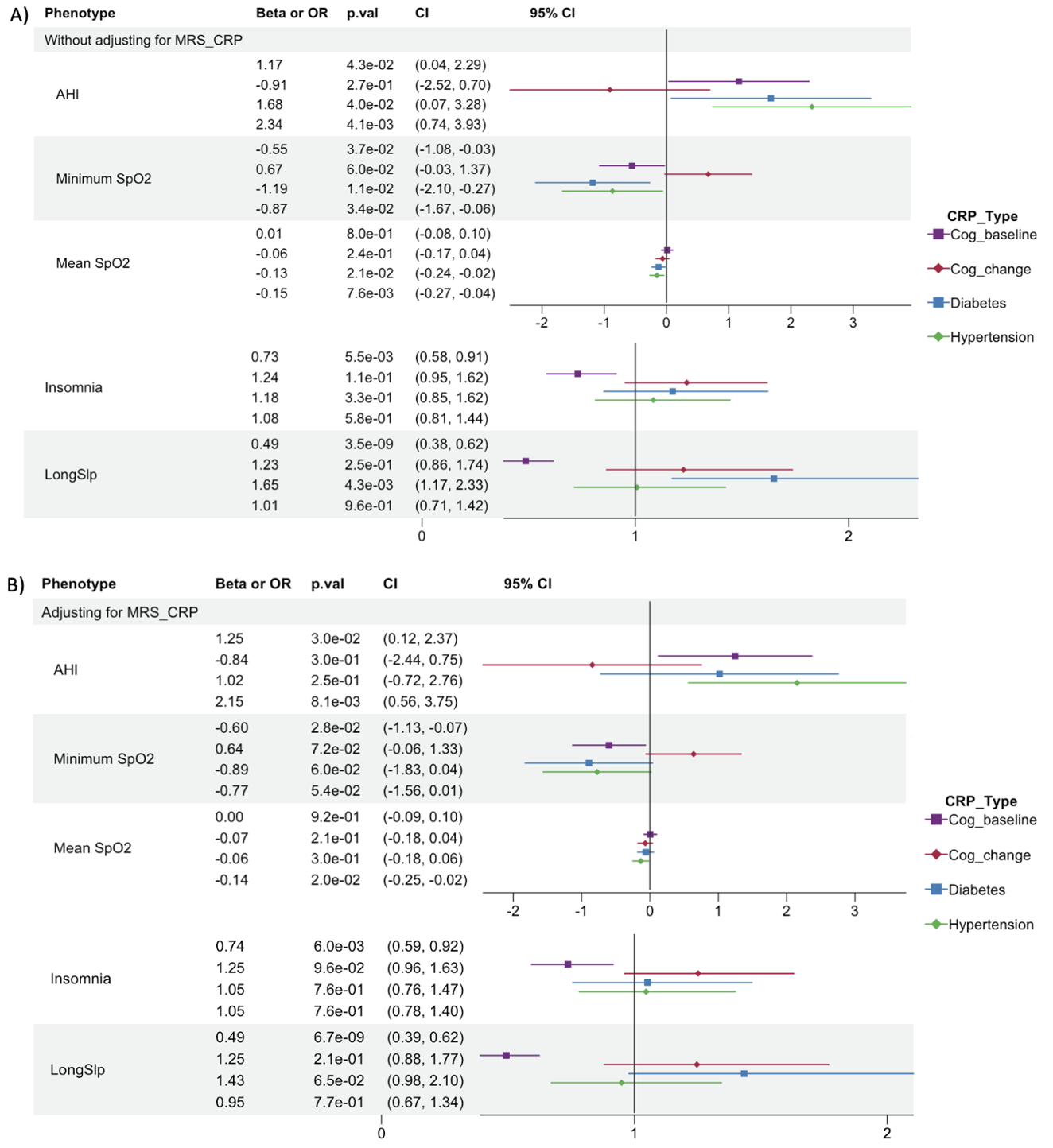

A) Association results without adjusting for MRS-CRP; B) Association results with MRS-CRP adjusted as covariate in the model. From left to right model coefficients (Beta or OR: odds ratio), p value (p.val) and 95% confidence interval (CI). AHI: apnea hypopnea index; SpO2: oxygen saturation; Cog_baseline: cognitive function score at baseline; Cog_change: change in cognitive function score between baseline and SOL-INCA visit; LongSlp: long sleep (sleep duration > 9 hours).

#### Figure S4. Forest plot comparing associations between methylation risk score for C-reactive protein (MRS-CRP) and health outcomes with and without adjusting for polygenic risk score (PRS) for CRP in HCHS/SOL.

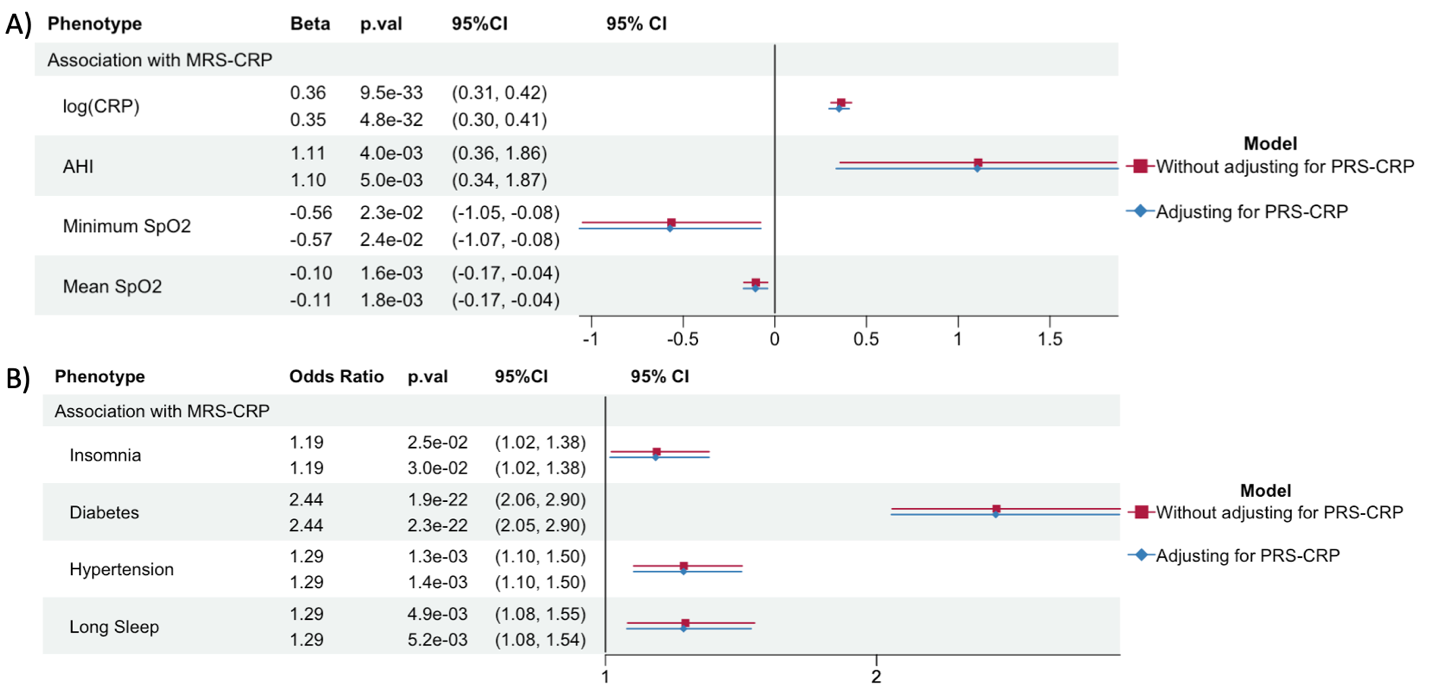

A) Association between MRS-CRP and listed phenotypes modeled as continuous variables; B) Association between MRS-CRP and listed phenotypes modeled as binary variables. From left to right model coefficients or odds ratio in case of binary outcomes, p value (p.val) and 95% confidence interval (95% CI). Log(CRP): log-transformed blood CRP level; AHI: apnea hypopnea index; SpO2: oxygen saturation.

#### Figure S5. Cubic spline model fit plot for methylation risk score for C-reactive protein (MRS-CRP) and blood CRP in HCHS/SOL

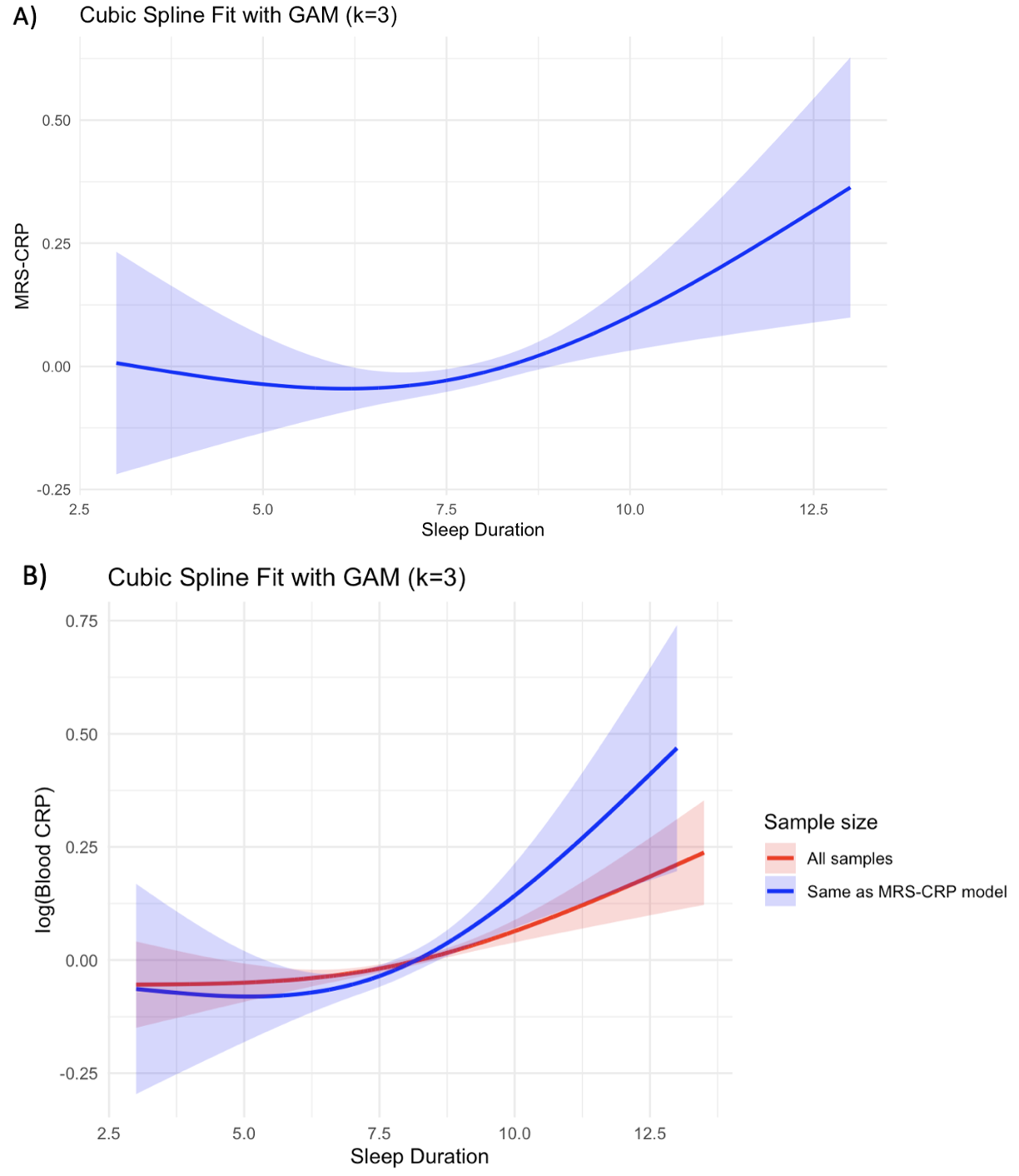

A) Cubic spline model fit for MRS-CRP; B) Cubic spline model fit for blood CRP level using all participants and same participants as MRS-CRP model.

Sleep Duration is measured in hours.

#### Figure S6. Forest plot showing associations between blood and polygenic risk score (PRS) for C-reactive protein (CRP) and health outcomes using all available HCHS/SOL participants

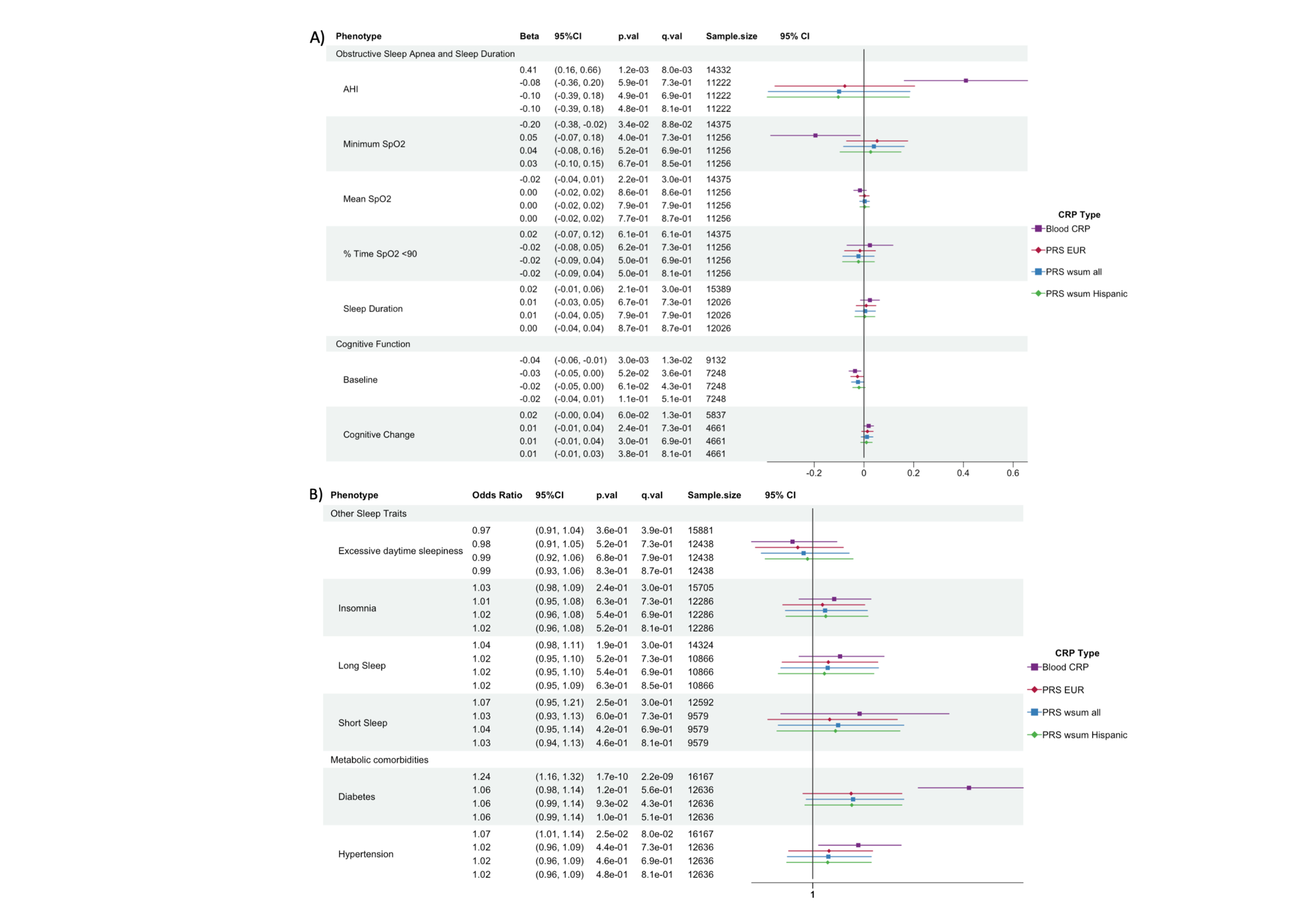

A) obstructive sleep apnea (OSA), sleep duration and cognitive traits; B) odds ratio for binary cardio-metabolic and other sleep outcomes. From left to right model coefficients or odds ratio in case of binary outcomes (Beta), 95% confidence interval (95% CI), p value (p.val), FDR corrected q value (q.val) and sample size.

#### Figure S7. Forest plot comparing effect size of the methylation risk score for C-reactive protein (MRS-hsCRP) associations of model 1 and sensitivity analyses in HCHS/SOL.

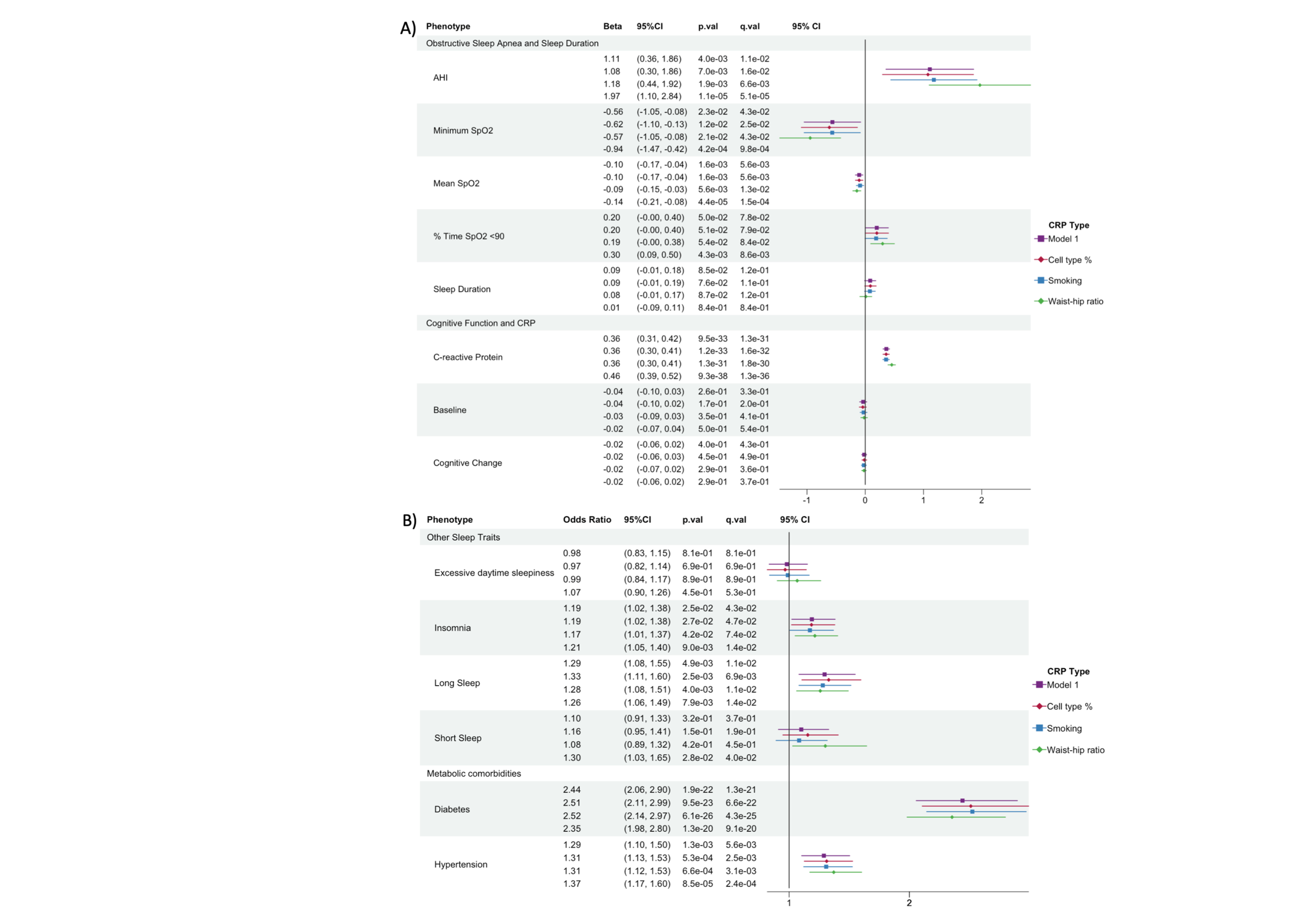

A) obstructive sleep apnea (OSA), sleep duration and cognitive traits; B) odds ratio for binary cardio-metabolic and other sleep outcomes. From left to right model coefficients or odds ratio in case of binary outcomes, p value (p.val), FDR corrected q value (q.val) and 95% confidence interval (95% CI). AHI: apnea hypopnea index; SpO2: oxygen saturation; Cognitive function: change in cognitive function score between baseline and SOL-INCA visit (Change); cognitive function score at baseline (Baseline).
